# Critical levels of mask efficiency and of mask adoption that theoretically extinguish respiratory virus epidemics

**DOI:** 10.1101/2020.05.09.20096644

**Authors:** Alan D. Kot

## Abstract

Using a respiratory virus epidemiological model we derive equations for the critical levels of mask efficiency (fraction blocked) and of mask adoption (fraction of population wearing masks) that lower the effective reproduction number to unity. The model extends a basic epidemiological model and assumes that a specified fraction of a population dons masks at a given initial number of infections. The model includes a contribution from the ocular (nasolacrimal duct) route, and does not include contributions from contact (fomite) routes. The model accommodates dose-response (probability of infection) functions that are linear or non-linear. Our motivation to study near-population-wide mask wearing arises from the concept that, between two mask wearers, the concentration of particles at inhalation should be the square of the mask penetration fraction. This combination, or team, of masks can provide a strong dose-lowering squaring effect, which enables the use of lower-efficiency, lower-cost, lower pressure-drop (easier breathing) masks.

For an epidemic with basic reproduction number *R*_0_ = 2.5 and with a linear dose-response, the critical mask efficiency is calculated to be 0.5 for a mask adoption level of 0.8 of the population. Importantly, this efficiency is well below that of a N95 mask, and well above that of some fabric masks. Numerical solutions of the model at near-critical levels of mask efficiency and mask adoption demonstrate avoidance of epidemics. To be conservative we use mask efficiencies measured with the most-penetrating viral-particle sizes. The critical mask adoption level for surgical masks with an efficiency of 0.58 is computed to be 0.73. With surgical masks (or equally efficient substitutes) and 80% and 90% adoption levels, respiratory epidemics with *R*_0_ of about 3 and 4, respectively, would be theoretically extinguished.

## 1. Introduction

When a novel respiratory virus epidemic begins exponential growth in a population, its novelty implies that there is no community (herd) immunity, nor a vaccine. The population can undertake various non-pharmaceutical measures such as physical distancing, quarantining (based on symptoms, testing, contact-tracing), work and school closures, travel restrictions, careful hand-hygiene and wearing masks (Ferguson et al., 2006, 2020). While applying all of these measures simultaneously would be the most effective, the impact on society may become immense. Therefore it is valuable to gauge the benefit provided by various measures. Herein we use a mathematical epidemiological model to examine the efficacy of near population-wide adoption of masks.

Our motivation to examine wide adoption of masks arises from the concept that between two mask wearers, the concentration of particles at inhalation (after exhalation and normalization to unity) should be the square of the mask penetration (fraction transmitted). (This concept is outlined in Figure 1.) Since squaring can be a much more powerful effect than a linear effect, some significant gains may remain for less efficient masks and incomplete adoption by the population. This combination, or team, of masks may provide a strong dose-lowering squaring effect, which would enable the use of lower-efficiency, lower-cost, lower pressure-drop (easier breathing) masks.

**Figure 1:**
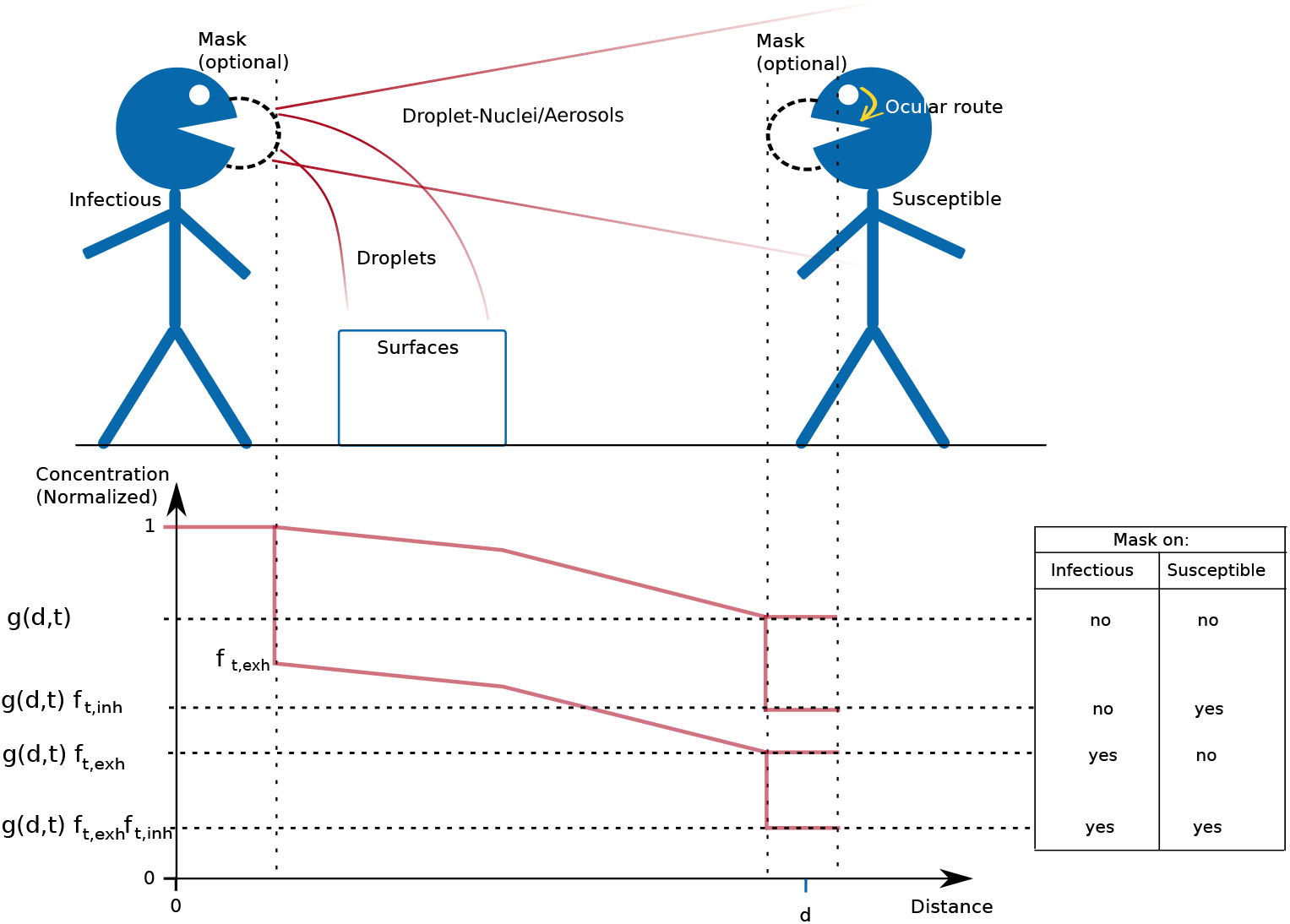
Routes of Transmission and Normalized Concentration versus Distance. The upper half of the diagram represents routes of transmission. The lower half of the diagram represents the concentration of particles (normalized at the source, and along a straight line between faces). The non-normalized concentration would depend on the event, e.g. exhalation, speaking, coughing, sneezing. The gain *g*(*d, t*) ≤ 1 is a non-linear function of distance, time, the event, and other factors. For sufficient distance the concentration would decline such that infection becomes unlikely without masks. The mask on the infectious person is modelled as a filter with a transmission gain (fraction that penetrates) during exhalation of 0 < *f*_*t,exh*_ ≤ 1. Similarly, the mask on the susceptible person has a filter transmission during inhalation of 0 < *f*_*t,inh*_ ≤ 1. Not wearing a mask corresponds to a filter transmission of 1. The four combinations of mask wearing, (none, either one, both) lead to four lower concentrations given by the respective products of the mask filter gains. For identical masks with low-flow events (e.g. breathing, speaking) the exhalation and inhalation filter gains may be modelled as being equal, *f*_*t*_. In that case, with both people wearing masks, the product of the filter gains is the square of their values. For example, when both people wear a *f*_*t*_ = 0.1 mask, then the resultant concentration 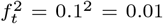, which is 100 times lower than no masks, and 10 times lower than a single mask. While this advantage decreases for weaker filters, the potential strength of that squaring effect motivates examination of what mask filtering ability and mask adoption level would significantly reduce epidemic strength. The ocular route is through the nasolacrimal duct and its significance is discussed in the text. For influenza, it appears to be on the order of 100 times less significant than inhalation.

Respiratory virus transmission can occur via droplet, aerosol (including droplet nuclei formed by evaporation of small droplets) and contact routes (Atkinson and Wein, 2008; Wei and Li, 2016; Xie et al., 2007). High-speed photography of visible droplets during sneezing events have shown ranges of roughly 8m (Bourouiba, 2020), but such events are rare (Atkinson and Wein, 2008; Wein and Atkinson, 2009), especially during a pandemic where generally people have a heightened awareness to not cough or sneeze among others. High-speed laser-aided photography of talking at close-range revealed numerous visible droplets in the range of 20-500 *µ*m (Anfinrud et al., 2020; Stadnytskyi et al., 2020). Droplets captured at close-range and smaller than 5 *µ*m have been reported to contain about nine-fold more viral copies than larger particles (Milton et al., 2013). Small droplets can evaporate to form droplet-nuclei that can remain airborne (Xie et al., 2007; Stadnytskyi et al., 2020). Talking for five minutes can generate the same number of droplet nuclei as a cough (Xie et al., 2007). The viability of aerosol viral particles has been demonstrated for SARS-CoV-1 and SARS-CoV-2 for the three hour duration of an experiment (van Doremalen et al., 2020), and for 16 hours in (Fears et al., 2020). Non-symptomatic and pre-symptomatic transmission can be significant (Wei et al., 2020). An example of this is where 40 of 60 participants at a choir practice became infected with COVID-19, despite distancing and without coughing or sneezing (Wei et al., 2020). With SARS, community wearing of masks when going out had a significant protective odds ratio of 0.36 compared with not wearing a mask (Lau et al., 2004). Mask wearing can reduce exposure especially in situations where physical distancing is difficult. Taiwan, with a population of 24 million, has only had seven COVID-19 deaths, had significant surgical mask use plus other measures and early action (Taylor et al., 2020).

While many masks, including home made fabric masks, might be efficient at blocking droplets, we take a more conservative approach and use mask efficiencies for aerosol sized particles of about 0.02-1 *µ*m. Thus, if such masks were stored by a population in advance of a novel respiratory virus, their rated filtering would apply to a wide range of particle sizes and regardless of whether the novel virus is transmitted predominantly by droplets or aerosols.

Various prior works have investigated epidemiological modelling of respiratory viruses with masks, including (Atkinson and Wein, 2008; Brienen et al., 2010; Cui et al., 2019; Myers et al., 2016; Stilianakis and Drossinos, 2010; Tracht et al., 2010; Wein and Atkinson, 2009; Yan et al., 2019). In a pair of papers (Atkinson and Wein, 2008; Wein and Atkinson, 2009) Atkinson and Wein construct detailed concentration-based models of aerosol transmission of influenza with an epidemic model for households. In (Brienen et al., 2010) a simplified effect from mask usage is assumed. Tracht et al. (2010) used a detailed SEIR model for H1N1 influenza and demonstrated various degrees of effect, including extinguishing the epidemic in some cases. Cui et al. (2019) added compartments to include asymptomatic transmission. Yan et al. (2019) used a detailed concentration-based model for Influenza. Some recent works provide detailed models for Covid-19, including (Eikenberry et al., 2020; Ngonghala et al., 2020) and demonstrate the efficacy of masks and other interventions.

Herein we determine closed-form expressions for the critical mask efficiencies and mask adoption levels that can extinguish a respiratory virus epidemic, including linear or non-linear probability of infection (dose-response) functions, and including contributions from the ocular (nasolacrimal) route. We use a relatively simple model, which requires few parameters and is consequently easier to apply to a novel virus. With this simpler model, in some cases one needs only the fundamental parameter *R*_0_.

## 2. Methods

### 2.1. Transmission Routes

Figure 1 gives a rough overview of the various routes of transmission from an infectious person to a susceptible person.

The lower half of Figure 1 contains a plot that represents the concentration of airborne particles versus distance (not to scale). It is intended to give a conceptual overview of the normalized concentration of virulent particles along a line between the infectious’ face and a susceptible’s face. The non-normalized concentration would depend on the event (e.g. exhalation, speaking) and on the local environment.

The decrease in concentration with distance is shown in Figure 1 as a gain, *g*(*d, t*) ≤1, being a non-linear function that typically decreases with distance, and depends on time, and on other factors such as the event, humidity, and temperature. Here we consider only how masks affect the concentration in a relative manner, without needing to model the complicated function *g*(*d, t*). Note that for low-volume events (e.g. breathing, talking) that the region of higher concentrations would be less like the cone shown in Figure 1, and a more like a cloud nearer the source (Stilianakis and Drossinos, 2010).

### 2.2. Dose Lowering by Masks

In Figure 1 the mask on the infectious person is modelled as a filter with a transmission gain (fraction that penetrates) during exhalation of 0 < *f*_*t,exh*_ ≤ 1. Similarly, the mask on the susceptible person has a filter transmission during inhalation of 0 < *f*_*t,inh*_ ≤ 1. Not wearing a mask corresponds to a filter transmission of unity. The four combinations of mask wearing, (i.e none, either one, or both) are shown to lead to lower concentrations given by the product of the mask filter gains.

The dose input to the respiratory tract may be modelled as an integral over time of the concentration at the susceptible’s face,

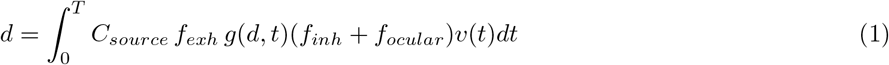

where *C*_*source*_ is the concentration at the source, and *v*(*t*) is the volume of air inhaled. (The contribution for the ocular route is discussed in the next section.)

We take the filter properties to be constants (either during low-flow events like breathing, or use averages for more dynamic events). Consequently,

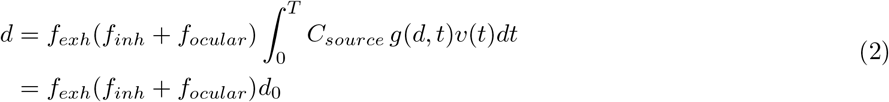

so that the filter gain *f*_*exh*_(*f*_*inh*_ + *f*_*ocular*_) < 1 reduces the original dose *d*_0_ by a set of four constants, where the values of *f*_*exh*_ or *f*_*inh*_ are set to unity for those not wearing a mask.

### 2.3. The Ocular Route

The ocular route corresponds to collection of particles by the eye which then pass through the nasolacrimal duct (Bischoff et al., 2011). The ocular route for viral particles is modelled in Figure 1 as a passage of the incident concentration through a filter denoted *f*_*ocular*_. It is implicit in the dose integral above that *f*_*ocular*_ was measured during breathing. We estimate *f*_*ocular*_ based on data in Figure 2 of (Bischoff et al., 2011), as follows. The control group (Group 1) in (Bischoff et al., 2011) had no masks or goggles, and an ocular-exposure-only group (Group 2) had a half-mask with clean air supply. These groups were equally exposed to a standardized aerosol concentration of live attenuated influenza vaccine. Afterwards they used nasal washes followed by quantitative reverse transcription polymerase chain reaction (RT-PCR) to count the resulting number of Influenza RNA copies. The control group would have had count-contributions from both inhalation, *n*_*inh*_, and from ocular paths, *n*_*oc*_, so its count of 504 consists of *n*_*inh*_ + *n*_*oc*_. The ocular-path-only group count had *n*_*oc*_ = 5. Assuming linearity of the quantitative RT-PCR tests, then the ratio *n*_*oc*_/*n*_*inh*_ = 0.01 is the fraction of viral particles that pass through the ocular route compared to the inhaled route, which is indeed the parameter *f*_*ocular*_ that we seek. The ocular route transmission is about 100 times lower than inhaling without a mask, so masks would be the most effective first-approach to lowering the dose.

**Figure 2:**
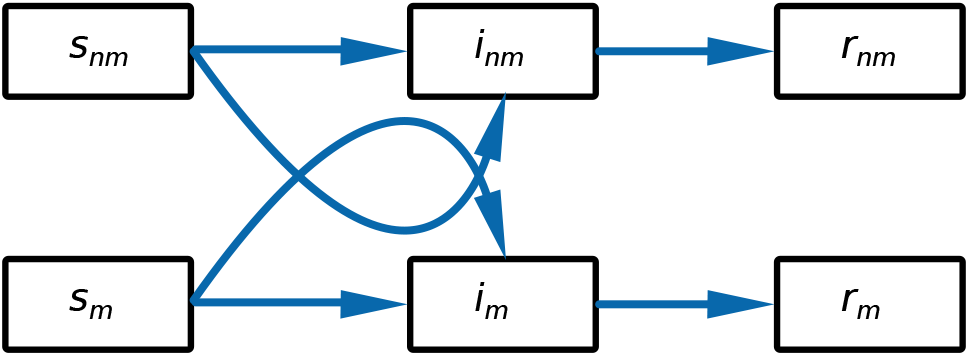
SIR Model of Respiratory Epidemic with Mask Wearing. The upper row of compartments accounts for those with no mask and the lower row accounts for those wearing a mask. The curved line from *s*_*m*_ to *i*_*m*_ passes near *i*_*nm*_ to graphically indicate infection of masked by non-masked. Similarly, the curved line from *s*_*nm*_ to *i*_*nm*_ passes near *i*_*m*_ to graphically indicate potential infection of the non-masked by the masked.

Since *f*_*ocular*_ summarizes aerosol reception, but not droplets, direct exposure to droplets from coughs or sneezes would be sensibly reduced by eye protection (glasses, shields). In (Atkinson and Wein, 2008) the occurrence of cough and sneezes was modelled to be rare. Moreover, in the context of a pandemic, people have a heightened awareness of the need not to cough or sneeze in close to others, so the contribution from coughing and sneezing would be reduced.

With more potent viruses or in environments of higher exposure, higher efficiency masks would be used, in which case their transmission fraction lowers to nearer that of the ocular route, so that the ocular route becomes significant. In such situations the use of sealed goggles or similar should be considered.

The ocular filter value estimate of 0.01 is low relative to typical mask transmission in our examples. It will be seen later in the numerical results that the contribution from the ocular route barely affects the critical mask efficiency.

### 2.4. The Contact Route

The contact route was not included for several reasons. First, it’s been estimated to not be significant compared to the droplet/aerosol route (Atkinson and Wein, 2008). Second, during an epidemic there is heightened awareness of not touching one’s face without hand washing. Third, mask wearing among a significant proportion of the population would decrease viral deposition on fomites. Fourth, mask wearing impedes the ability to touch one’s nose or mouth. Fifth, the median-effective dose for influenza via the oral route is about 500 times higher than the nasal route (Atkinson and Wein, 2008). All of these assumptions may not be valid other viruses or for subsets of the population, such as children. For children, one would hope that children too young to learn the value of hand washing are well supervised and in clean environments, and that ones that are old enough to learn are well taught.

### 2.5. Probability of Infection

The effect of mask filtering is to lower the dose, and the effect of that lower dose on probability of infection is given by a dose-response function, denoted *p*_*i*_ = *P*_*i*_(*d*).

In common SIR epidemiological models recall that the parameter *β* is the expected number of contacts per day that cause infection (Hethcote, 2000). It is the expected number of infections per day from those contacts, or

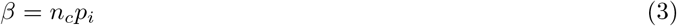

where *n*_*c*_ is the expected number of contacts per day, and *p*_*i*_ is the probability that such a contact results in an infection.

To consider how lowering the dose affects the probability of infection, let *p*_*i*0_ denote the probability of infection for the no-mask situation. Since *p*_*i*0_ corresponds to some dose *d*_0_, we can find *d*_0_ from

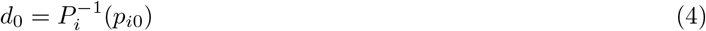

where 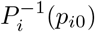 denotes the inverse of the dose-response function *P*_*i*_(*d*).

For a modification of the dose by multiplying it by some value *m*, then the resulting probability of infection is

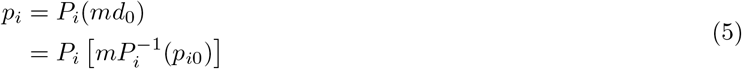

For cases where the dose response function is linear and *p*_*i*0_ corresponds to a dose in the linear region, then

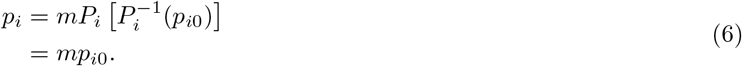

Consequently, for mask use with a linear dose-response function and linear no-mask-dose, *m* = *f*_*exh*_(*f*_*inh*_+*f*_*ocular*_) < 1 then

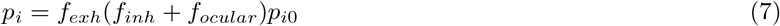

so that *p*_*i*_ is scaled down linearly by the filtering.

For the case where *f*_*ocular*_ is insignificant and the mask transmissions are identical

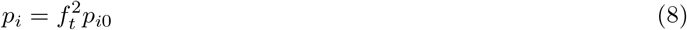

so the probability of infection is lowered by the square of the filter transmissions. This squaring effect is potentially stronger for higher efficiency filters.

In addition to the linear dose-response case, we will later consider exponential, and approximate beta Poisson dose-response functions.

### 2.6. Mask Efficiency

Before describing the epidemiological model, we discuss some assumptions on mask efficiencies. In a variety of work on masks, including (Davies et al., 2013; Jung et al., 2014; Lee et al., 2008; Leung et al., 2020; Milton et al., 2013; Mueller and Fernandez, 2020; Oberg and Brosseau, 2008; Rengasamy et al., 2010; Smith et al., 2016; van der Sande et al., 2008) there are variations in measured efficiencies arising from various testing procedures, particle sizes, and fit. Masks are typically much better at blocking larger droplets, therefore it is prudent to measure mask efficiency at the most-penetrating particle-size (MPS) for infective particles. Viral particles and bacteria range from about 0.02 to 1.26 *µ*m (Lee et al., 2008). (It was noted in (Lee et al., 2008) that the size of the influenza and SARS-CoV-1 viruses happen to coincide with the most-penetrating particle-size for N95 respirators.) If a population stocked MPS-rated masks in advance of a novel respiratory virus, then the masks would meet or exceed those worst-case efficiencies over a wide range of particle sizes.

Herein we do not attempt to compare a wide variety of masks, since our principal aim is the critical mask efficiency for extinguishing an epidemic. However in some numerical examples we assume a surgical mask. Based on (Lee et al., 2008) the MPS efficiency of a surgical mask is taken to be 0.58. That efficiency is comparable to that in (Milton et al., 2013), where the reduction in quantitative RT-PCR viral counts from fine (less than 5 *µ*m) particles, after exhalation through a surgical mask, was 2.8 fold (which corresponds to an efficiency of 0.64). Over all particle sizes the effect of the surgical mask was a 3.4 fold reduction (which corresponds to a 0.7 efficiency) (Milton et al., 2013). Some fabrics are very poor viral filters with efficiencies of roughly 0.1 (Rengasamy et al., 2010).

Later, for some equations we will assume that the exhalation and inhalation filter transmission gains are equal. Near equal penetration for inhalation and exhalation using surgical masks was reported in (Jung et al., 2014). Note also that the inhalation mask efficiency of 0.58 in(Lee et al., 2008) was comparable to the exhalation mask efficiency of 0.64 in (Milton et al., 2013). Equal filtering in either direction may be reasonable for low-flow events like breathing and talking, and perhaps less reasonable for high-flow, but rare, events like coughing and sneezing, depending on the mask seal and construction.

A common concern about surgical and other types of masks, including home-made masks, is leakage between the mask and face. A significant and low cost method to improve fit is an overlay of a nylon stocking (Cooper et al., 1983; Mueller and Fernandez, 2020). As reported in (Mueller and Fernandez, 2020) the use of a nylon stocking overlay raised the efficiency of five of ten fabric masks above a benchmark surgical mask.

### 2.7. SIR Compartment Model

A basic mathematical model of an epidemic is the SIR (Susceptible, Infectious, Recovered) model (Hethcote, 2000). Individuals in those categories are modelled as transitioning from one compartment to the next according to various rate parameters, with a corresponding set of simultaneous differential equations.

We will use extensions of the SIR model to include mask use and will focus on the number of peak infectious and the final cumulative infected. All recovered are assumed to have immunity.

The basic SIR model is often augmented with an exposed compartment to form a SEIR model. However, note that every person accounted for through the I, or the R, compartments do so in both the SIR and in the SEIR models. Consequently, for identical corresponding parameters, both the SIR and SEIR models will have the same value of final cumulative infected. The incubation period of the exposed compartment spreads the same flow of infections over a longer time period. Since herein we are interested in filter conditions for which an epidemic is extinguished, and since such conditions will be equivalent for the SIR and the SEIR model, then for our purpose it is simpler, sufficient, and more general to use a SIR model.

Figure 2 shows a SIR compartment model for a respiratory viral epidemic where a portion of the population wear masks. The upper row of compartments accounts for those with no mask and the lower row accounts for those wearing a mask.

For convenience we define the state variables to be fractions of the population, so that

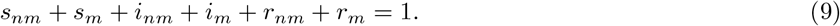

(e.g. *i* = *I*/*N* where *I* is the number of Infectious and *N* is the population.)

We also express the totals of Susceptible, Infectious, and Removed as *s* = *s*_*nm*_+*s*_*m*_, *i* = *i*_*nm*_+*i*_*m*_ and *r* = *r*_*nm*_+*r*_*m*_ respectively.

Initially none of the population is assumed to wear masks. When the beginning of an epidemic is detected by observing some specified number *i*_0_ of infections, then a specified fraction *p*_*m*_ of the population dons masks. These initial conditions are,

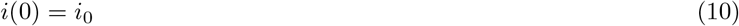

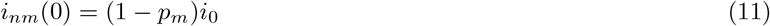

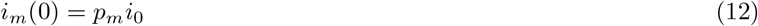

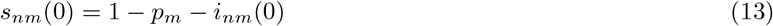

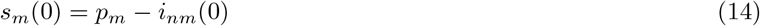

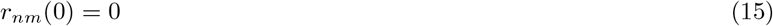

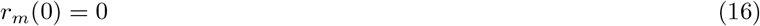

The set of differential equations for the model are,

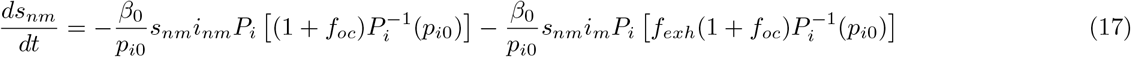

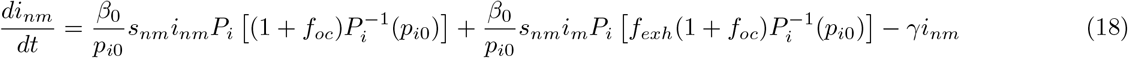

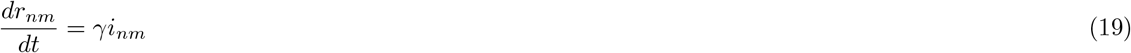

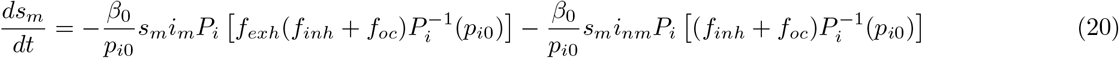

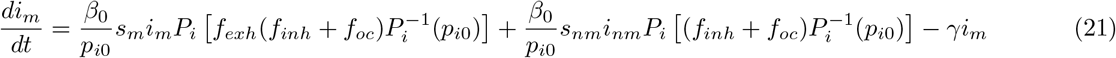

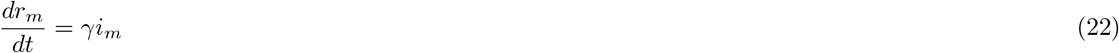

where;

*β*_0_ is the average number of contacts per day that are sufficient to cause infection, where the subscript 0 indicates no interventions. *β*_0_ is related to *β* (our case with interventions that affect the probability of infection) by

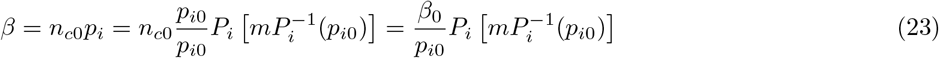

where the term 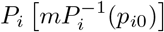 was discussed in Section 2.5 on the probability of infection, and where *m* is the dose-altering fraction. Note that *β* varies across the quartet of filter values, and importantly becomes a lower force of infection between contacts that have better filtering.

*γ* is the rate that the infected become recovered, which is the inverse of the duration of the infectious period.

*f*_*exh*_ and *f*_*inh*_ are the mask transmission gains during exhalation and inhalation, respectively. It is common to specify a mask by its efficiency, which is the fraction that it blocks, rather than which it transmits, so its efficiency is *f*_*b*_ = 1 − *f*_*t*_.

An alternative model would combine the removed compartments in Figure 2, if they have identical rate of entry from the infected states, but keeping them separate enables separate accounting (for example, to demonstrate that non-mask wearers are more likely to get infected).

### 2.8. Basic Reproduction Number

*R*_0_ = *β*_0_/*γ* is the basic reproduction number (Hethcote, 2000) in the basic SIR model (i.e. without masks). Below we briefly summarize some key results using *R*_0_ because they are both fundamental and will be used alongside our mask model.

*R*_0_ can be sensibly interpreted as the average number of infections produced by a single infectious person during their infectious period (Hethcote, 2000). The rate of growth of the infectious compartment is

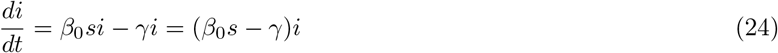

from which we can see that for the growth to be zero or negative, then *β*_0_*s* ≤ *γ*, or *β*_0_/*γ* = *R*_0_ ≤ 1. So, at the start of an epidemic, where *s* ≈ 1, there is no initial growth unless *R*_0_ > 1.

For community immunity, *R*_0_*s* < 1, or *s* < 1/*R*_0_. For example, for such immunity with *R*_0_ = 2.5 the susceptible fraction of the population must be less than 1/*R*_0_ = 1/2.5 = 0.4. Equivalently, the fraction of the population that has been through the infectious stage and has assumed immunity is 1 − 1/*R*_0_ = 0.6.

It is also possible to obtain analytical expressions for the peak infectious and the final cumulative infected (Hethcote, 2000), which are

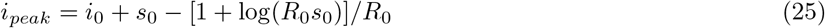

and *s*_∞_ the final value of *s*_*i*_, is given by the root of

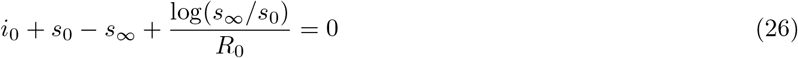

and *r*_∞_ = 1 − *s*_∞_.

## 3. Results

### 3.1. Effective Reproduction Number

For multi-compartment models van den Driessche and Watmough (2002) give the determination of the effective reproduction number *R*_0,*e*_. Of fundamental importance is that *R*_0,*e*_ = 1 is again a threshold, or a critical value, below which the epidemic is theoretically extinguished and above which growth will occur.

In Appendix A, using the methods of (van den Driessche and Watmough, 2002), *R*_0,*e*_ is found for the model of Figure 2 and its accompanying set of equations (10) - (22). The result is (44)

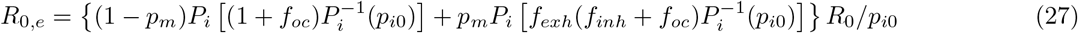

which can be simplified for identical mask transmissions, (45)

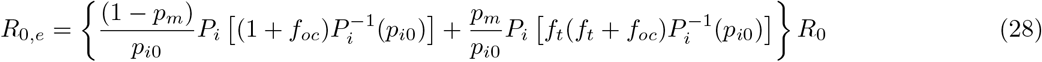

Additionally, if there is no ocular contribution, (46)

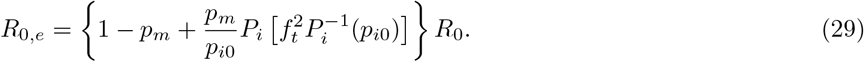

In Appendix B, we find *R*_0,*e*_ for linear, exponential, and approximate beta-Poisson dose response functions. A linear dose response is sometimes used in low-dose epidemiological models (Brouwer et al., 2017). An exponential dose-response has been used to model the infection probability of SARS-CoV-1 (Watanabe et al., 2010). Below we list some of the results from Appendix B.

#### 3.1.1. Linear Dose Response

From Appendix B equation (50), with a linear dose response, and identical filter transmissions *f*_*t*_,

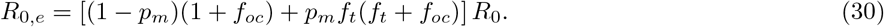

With no ocular contribution, (51)

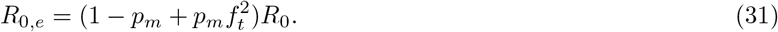

One can easily confirm that in the simple case of no masks, *p*_*m*_ = 0, then *R*_0,*e*_ = *R*_0_. Similarly, in the case of 100% mask adoption level, *p*_*m*_ = 1, then 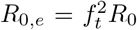 This result corresponds to our fundamental motivation described for Figure 1, where the result of transmission through two masks is the square of their individual transmissions.

In Figure 3 *R*_0,*e*_ given by (31) is plotted normalized (*R*_0,*e*_/*R*_0_) and using the mask efficiency *f*_*b*_ = 1 − *f*_*t*_. The linear dependence on *p*_*m*_ is clearer in Figure 3a. The non-linear dependence on *f*_*t*_, i.e the expected squaring effect of mask wearing, is clearer in Figure 3b.

**Figure 3:**
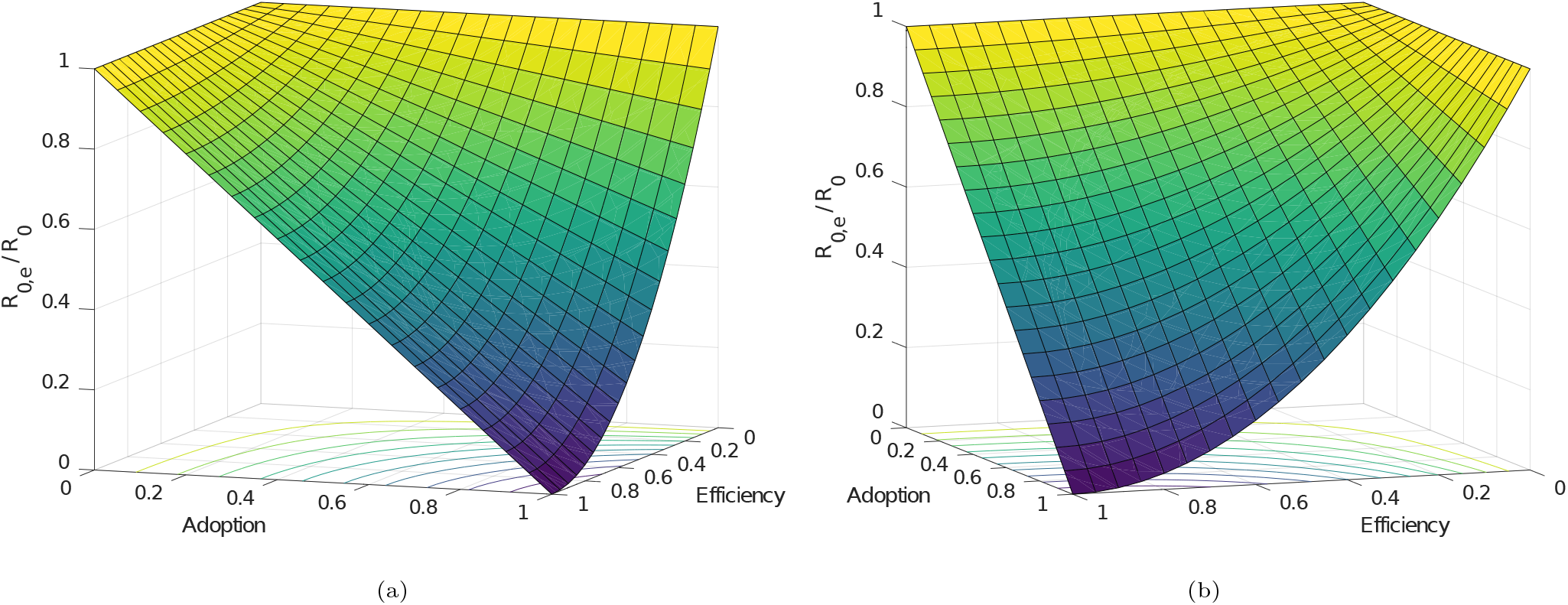
Normalized Effective *R*0 vs Mask Efficiency and Adoption. (Two views of the same surface.)

#### 3.1.2 Exponential Dose Response

From Appendix B equation (57), with an exponential dose response, with identical filter transmissions *f*_*t*_, and with no ocular contribution,

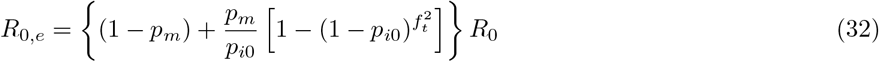

The more general cases of non-identical filters and including an ocular contribution can be found in Appendix B.

#### 3.1.3. Approximate Beta-Poisson Dose Response

From Appendix B equation (63), with an approximate beta-Poisson dose response, with identical filter transmissions *f*_*t*_ and with no ocular contribution,

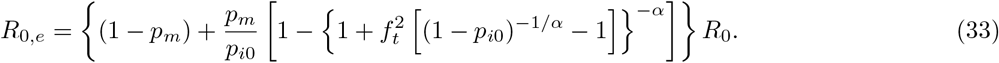

The more general cases of non-identical filters and including an ocular contribution can be found in Appendix B.

### 3.2. Critical Mask Efficiency

Here we find the filter efficiency for which *R*_0,*e*_ becomes equal to unity. In this section, we will assume masks with identical filter transmissions. In Appendix C, we find the critical mask efficiencies for linear, exponential, and approximate beta-Poisson dose response functions. Below we list some of those results.

#### 3.2.1. Linear Dose Response

With a linear dose response, from Appendix C (65)

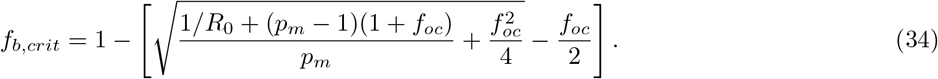

For no ocular contribution this simplifies to (66)

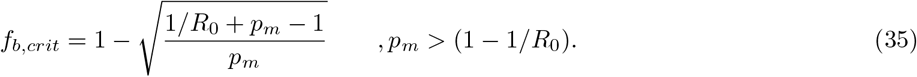

Figure 4a is a plot of the critical mask efficiency *f*_*b,crit*_ versus the mask adoption level *p*_*m*_, for various values of *R*_0_. Estimates of *R*_0_ for influenza is about 1.5 (Coburn et al., 2009), and for COVID-19 it is roughly 2.5 (Ferguson et al., 2020).

**Figure 4:**
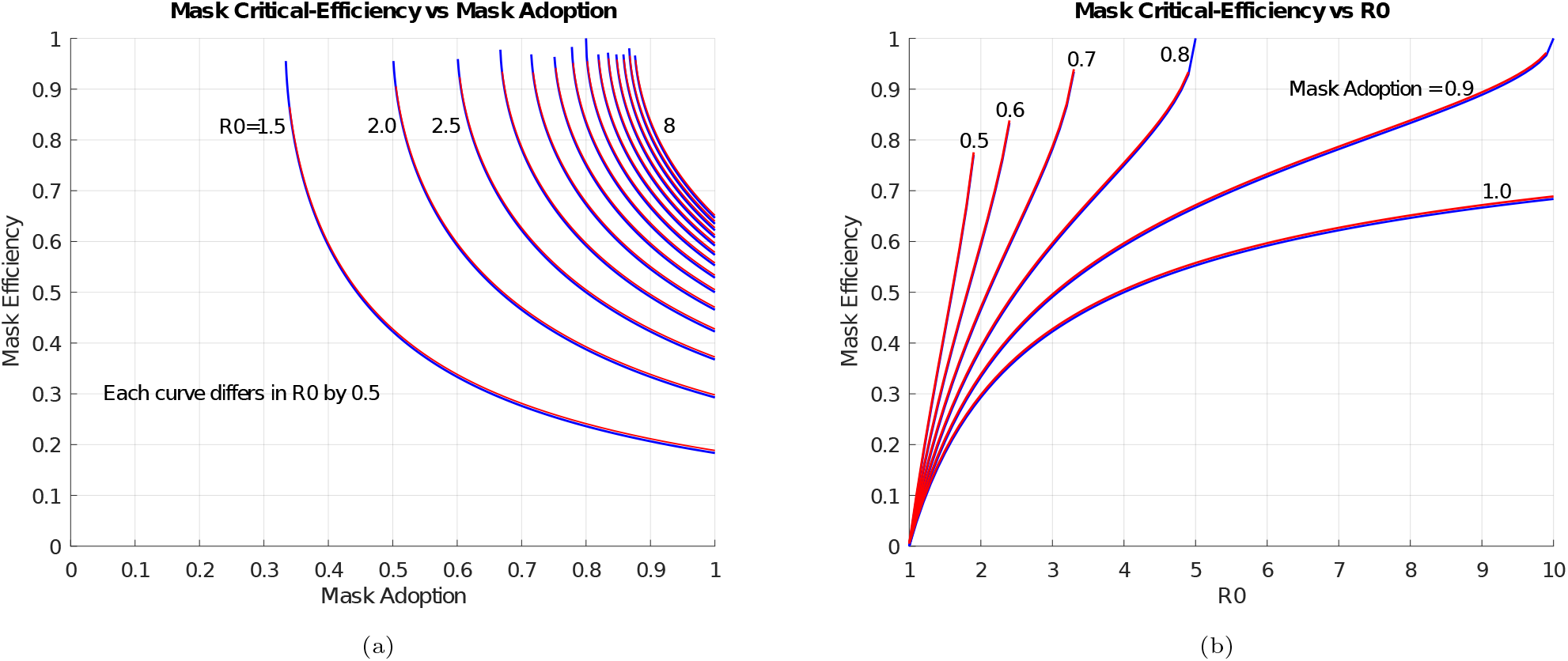
Mask Critical-Efficiency versus Mask Adoption and *R*_0_. Linear dose response.

As an example, assume that the mask adoption level is 80%, then from Figure 4a the *R*_0_ = 2.5 curve indicates a mask critical efficiency of 0.5. If the mask adoption rate was 90%, the mask critical efficiency is reduced to about 0.42. Importantly, these efficiencies are well below that of a N95 mask, and well above that of some fabric masks Rengasamy et al. (2010).

Figure 4b is a plot of the critical mask efficiency *f*_*b,crit*_ vs *R*_0_, for various values of mask adoption level *p*_*m*_. As *R*_0_ is increased, it can be seen that for a particular adoption level the required efficiency heads towards 100%, so after that even perfect masks would not suffice to extinguish the epidemic.

In both of Figures 4a and 4b, the upper curves (red) include the effect of the ocular route with *f*_*ocular*_ = 0.01. The curves with and without the ocular contribution (red and blue curves, respectively) are barely distinguishable. Accordingly we do not include the effect of the ocular route in examples in the remainder of this paper.

#### 3.2.2 Exponential Dose Response

With an exponential dose response, and neglecting the contribution for the ocular route, from Appendix C (69)

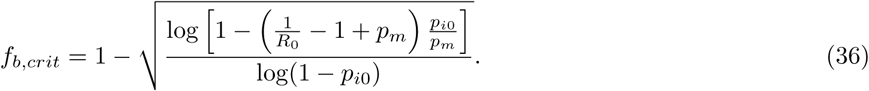

Figure 5a is a plot of the mask critical efficiency versus mask adoption for *R*_0_ = 1.5, 2.5, 3.5, for linear and exponential dose response functions. For the exponential case, we’ve shown both *p*_*i*0_ = 0.5 and *p*_*i*0_ = 0.25. Note that as *p*_*i*0_ is lowered the results are closer to the linear dose-response case. Recall that *p*_*i*0_ is the probability of a contact being infected, where this probability is representative of all contacts. Recall also that *R*_0_ = *n*_*c*0_*p*_*i*0_/*γ*, so that for example, with *R*_0_ = 2.5 and given an expected duration of infection 1/*γ* = 5, then *n*_*c*0_*p*_*i*0_ = *R*_0_*γ* = 2.5/5 = 0.5 and we can construct Table 1 with pairs that correspond to *n*_*c*0_*p*_*i*0_ = 0.5.

**Table 1:** Example pairs of *n*_*c*0_, *p*_*i*0_ whose product is 0.5.

|  |  |  |  |  |  |  |
| --- | --- | --- | --- | --- | --- | --- |
| $n_{c0}$ | 0.5 | 1 | 2 | 4 | 5 | 10 |
| $p_{i0}$ | 1 | 0.5 | 0.25 | 0.125 | 0.1 | 0.05 |

**Figure 5:**
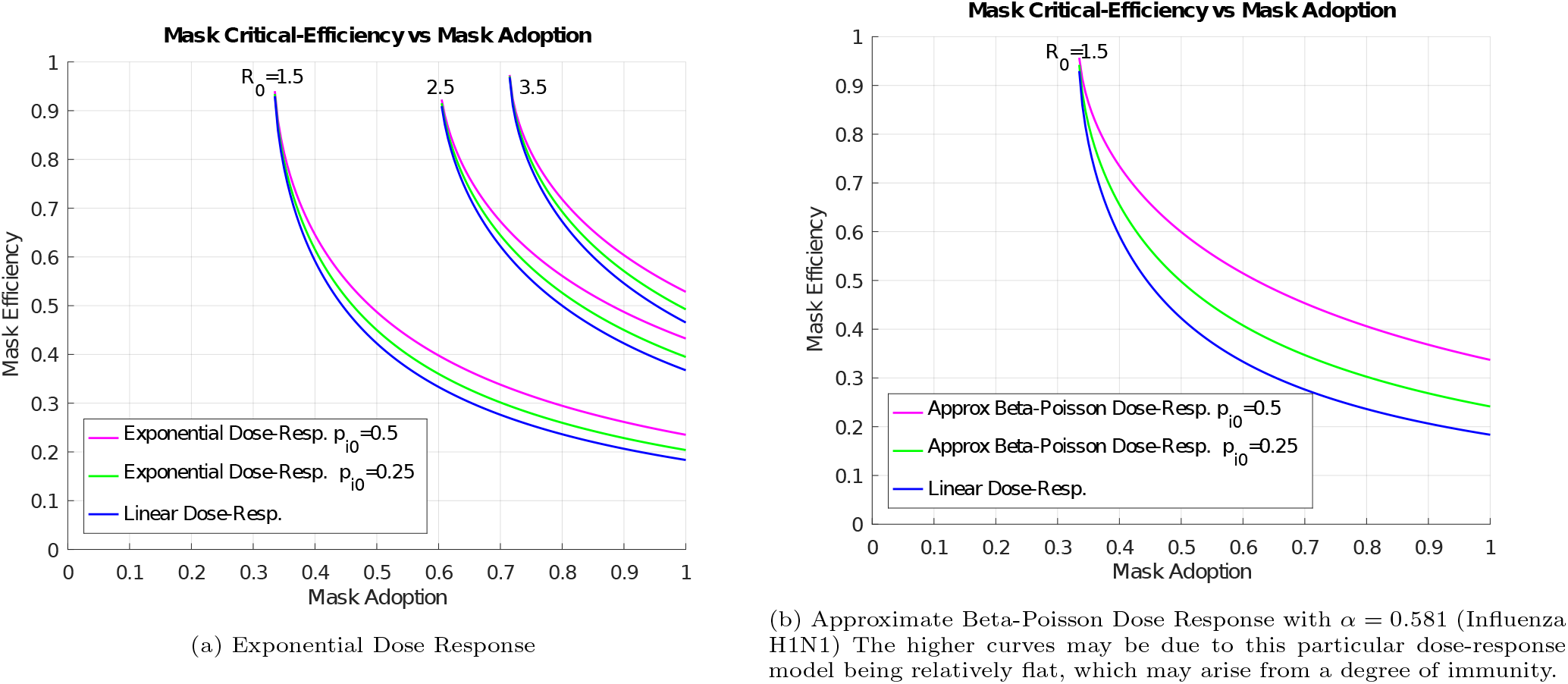
Mask Critical-Efficiency versus Mask Adoption, Non-Linear Dose Responses.

High values of *p*_*i*0_ imply near-certain transmission for each contact, which corresponds to doses high up the dose-response curve. In the absence of specific information on *p*_*i*0_ for a particular situation, one might use a median infective dose *p*_*i*0_ = 0.5 to represent the expected probability of infection. (This may not be correct in high-exposure situations, such as medical workers working closely with, and for several hours per day with infected patients.) For a given *R*_0_ a higher number of typical contacts implies a lower *p*_*i*0_. In (Watanabe et al., 2010) SARS-CoV-1 was well modelled by an exponential dose-response, and the estimated doses for residents of an apartment complex where an outbreak occurred were typically less than one-half of the median-infective dose. In (Mossong et al., 2008) the mean number of contacts per day was about 13 (although that contact count does not weigh the invectiveness of each contact, as is implicit in a SIR model).

#### 3.2.3. Approximate Beta-Poisson Response

With an approximate Beta-Poisson dose response, and neglecting the contribution for the ocular route, from Appendix C (72)

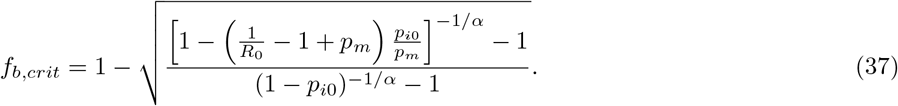

Figure 5b is a plot of the mask critical efficiency versus mask adoption for *R*_0_ = 1.5 for linear and approximate beta-Poisson dose response functions. The approximate beta-Poisson curves use the parameter *α* = 0.581 which models Influenza H1N1 (Murphy et al., 1984) with *R*_0,*e*_ = 1.5 (Coburn et al., 2009). With these parameters and for *p*_*i*0_ ≤ 0.25 Figure 5(b) shows that the critical mask efficiency is within about 25% of the linear dose-response result. With this value of *α* the dose-response curve is comparatively flat, requiring about 100 times the median-infective dose in order to infect 95% of the population – as if a portion of the population was immune. For the purpose of predicting the effects of masks in an epidemiological model, it would be preferable to have a dose-response curve where those with immunity are (somehow) not counted, and instead the immune portion are accounted for in the initial-value of the recovered.

### 3.3. Numerical Solution Examples

All of the examples in this section were produced by numerical solutions of the set of differential equations (10) - (22) for the model in Figure 2. Unless otherwise stated, all of these examples are for *R*_0_ = 2.5, an infectious duration of 5 days, an initial infected fraction of the population of 10^−4^, and with surgical masks taken to have an efficiency of 0.58.

The values used for *R*_0_ and for the infectious duration are roughly those for COVID-19 (Ferguson et al., 2020). Recall from the section describing the model, that for identical *R*_0_ and infectious duration, that both our SIR model or its extension to an SEIR model would have the same value of final cumulative infected.

Mainly we are interested in the overall results for the population, so we plot the totals of susceptible, infectious, and removed, i.e. *s* = *s*_*nm*_ + *s*_*m*_, *i* = *i*_*nm*_ + *i*_*m*_ and *r* = *r*_*nm*_ + *r*_*m*_ respectively.

**Figure 6:**
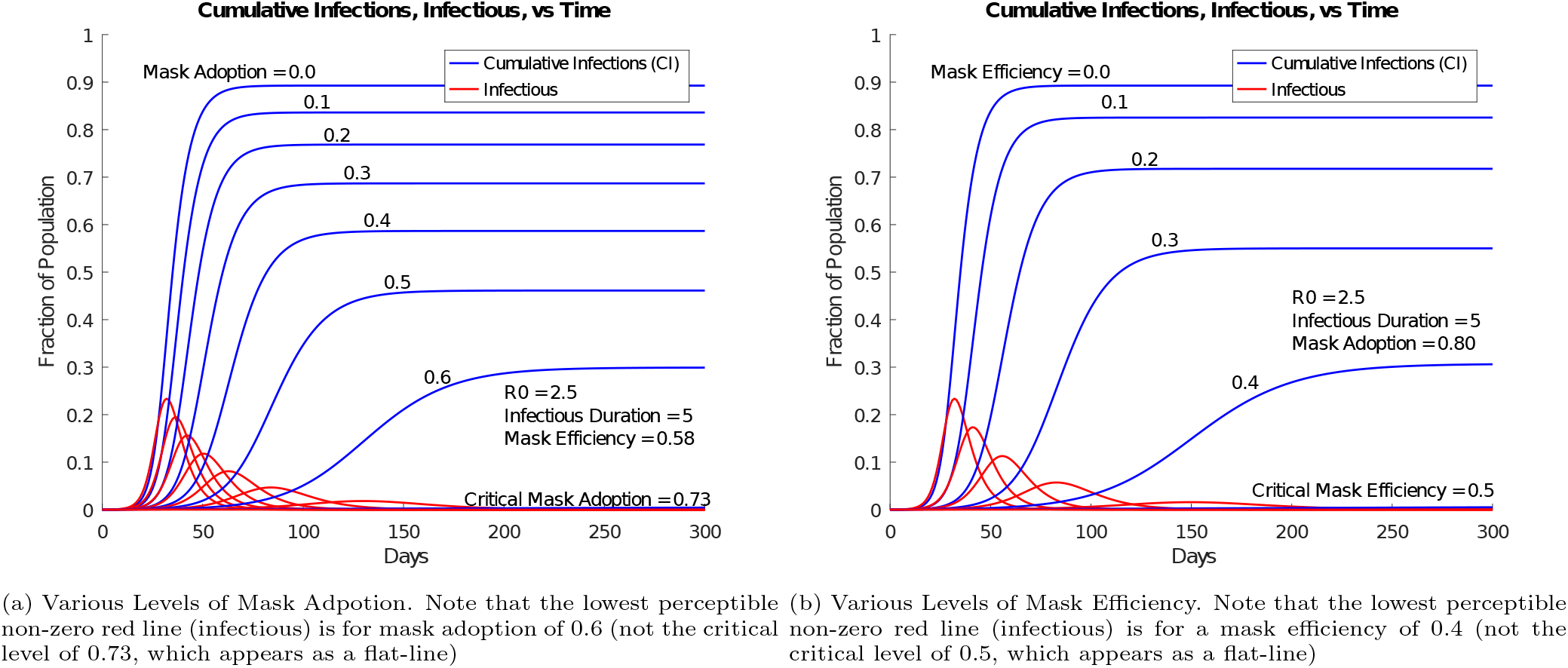
Infections vs Time, showing lowering epidemic as mask adoption (a) and mask efficiency (b) are increased towards their critical values. In each case, the critical levels appear as a flat-line. (In practice, one would aim for higher than critical values.)

Figure 6 shows plots of infections versus time, and shows lower curves as mask adoption and mask efficiency are increased towards their critical values.

Figure 6a shows lower curves as the mask adoption is increased towards its critical value of 0.73 (obtained from Figure 4a for a mask efficiency of 0.58 at *R*0 = 2.5). At the critical adoption level of 0.73, both the peak infected and cumulative infected curves appear to be flat at zero. Note that the lowest perceptible non-zero red line (infectious) on this plot is for a mask adoption of 0.6 (not the critical level of 0.73, which appears as a flat-line).

Figure 6b shows lower curves as the mask efficiency is increased towards its critical value of 0.5 (obtained from Figure 4a for a mask adoption of 0.8 at *R*0 = 2.5). At the critical efficiency of 0.5, both the peak infected and cumulative infected appear to be flat at zero. Note that the lowest perceptible non-zero red line (infectious) on this plot is for a mask efficiency of 0.4 (not the critical level of 0.5, which appears as a flat-line).

Figure 7 shows plots of log(infections) versus time, for two levels of mask adoption. The first level is at the critical adoption level and the second level is at a slightly higher-than-critical adoption.

**Figure 7:**
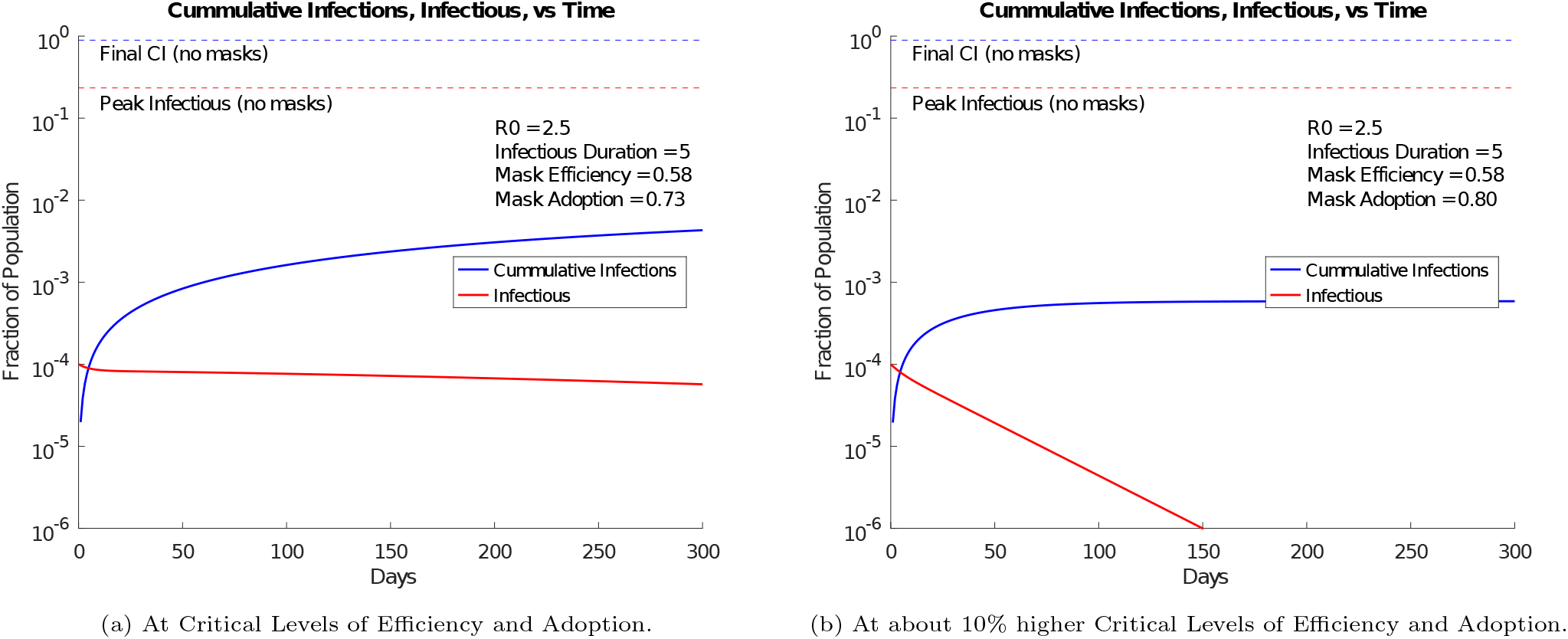
Log(Infections) vs Time.

Figure 7a is at the critical mask adoption level of 0.73. At that critical level, the infections are close to flat and near the initial value of 10^−4^ of the population. That low level of infections was too small to be visible on the non-log plot of Figure 6a.

Figure 7b is for a mask adoption level increased to 0.8 (about 10% over the critical value), and shows a clear extinguishing of the number of infectious.

Figure 8 shows linear and log plots of the *peak* infections and of the *final* cumulative infected, versus mask adoption, as the mask adoption is increased from zero to 100%.

**Figure 8:**
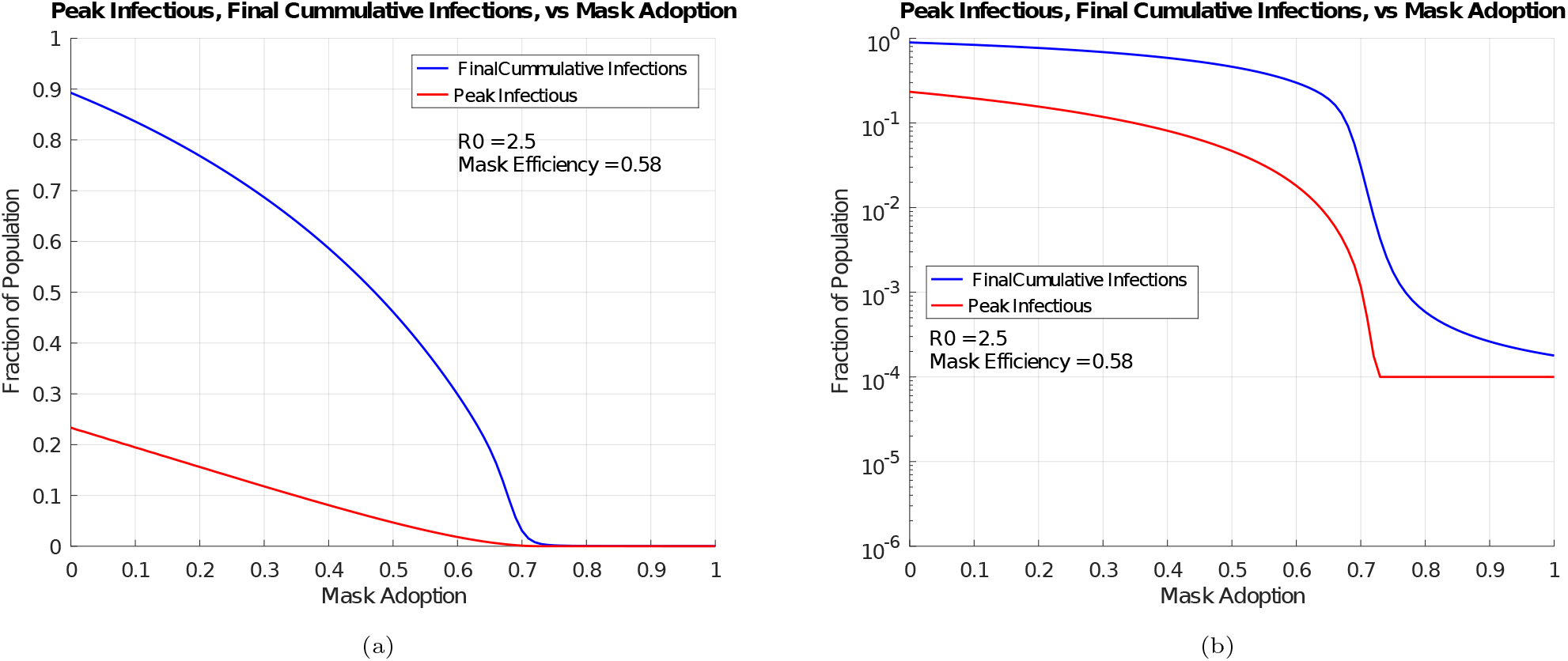
Infections vs Mask Adoption Level. Both (a) and (b) present the same data, but (b) is a log plot.

Figure 8a is a linear plot and demonstrates that the number of final cumulative infected has a sharp transition, or knee, near the critical level of mask adoption of 0.73.

Figure 8b is a log plot which more clearly shows a sharp transition near the critical level of mask adoption. The number of peak infectious drops to the number of the initial-infected as the critical value of mask adoption is approached.

## 4. Discussion

### 4.1. Summary

Equations were derived that give critical levels of mask efficiency and of mask adoption that lower the effective reproduction number *R*_0,*e*_ of a SIR-based epidemiological model to unity. Those critical levels correspond to a sharp reduction, or “knee”, towards zero in the final number of cumulative infected, as mask efficiency or mask adoption are increased. Consequently, rather than have a population wear masks of random or non-standardized efficiencies, it would be much more effective for the population to adopt masks that exceed the critical mask efficiencies derived herein.

The model used assumes that a specified fraction of a population dons masks at a given initial number of infections, and that there is no further influx of infectious people from outside the population.

The model does not include contributions from indirect-contact (fomite) routes, assuming that sufficient hand hygiene is achieved by the population, and assuming that broad mask adoption would decrease deposition onto fomites and would also decrease any tendency of mask wearers to touch their nose or mouth.

The model includes the ocular (nasolacrimal duct) route, and excludes direct contact from droplets. In our examples the effect on the critical mask efficiency from the ocular route was estimated to be insignificant. The ocular route would become more significant in environments of high exposure, where higher efficiency masks would be more suitable. With higher efficiency masks their transmission fraction lowers to nearer that of the ocular route, so that the ocular route may become significant.

The model accommodates dose-response (probability of infection) functions that are linear or non-linear. In our *R*_0_ = 2.5 example for an exponential dose-response, the increase in the critical mask efficiency was about 12% (assuming a probability of infection per contact being 0.5 with the number of contacts per day being 1). That increase drops to the linear case for a higher number of contacts per day.

Various assumptions were made in order to model the efficacy of population-wide mask wearing on its own. These assumptions include; no vaccinations, no physical distancing, no testing for being infectious (therefore no quarantining from testing), and symptom-less transmission (therefore no self quarantining). Such assumptions also enable a very simple and general model with few parameters. Indeed the simplest of the equations derived for the critical mask efficiency need only the fundamental parameter *R*_0_.

With the above model in an epidemic with *R*_0_ = 2.5 and with a linear dose-response, the critical mask efficiency is calculated to be 0.5 for a mask adoption level of 0.8 of the population. Importantly, this efficiency is well below that of a N95 mask, but well above that of some fabric masks.

To be conservative we use mask efficiencies reported for the most-penetrating particle-sizes. With *R*_0_ = 2.5, surgical masks with an efficiency of 0.58 give a computed critical mask adoption level of 0.73. With surgical masks (or equally efficient substitutes) and 80% and 90% adoption levels, respiratory epidemics with *R*_0_ of about 3, and 4, respectively, would be theoretically extinguished.

Numerical solutions of the model at near-critical levels of mask efficiency and mask adoption, for a 10^−4^ initially infected fraction of the population, demonstrate avoidance of epidemic growth (and so numerically confirm the equations derived for the critical levels). Example numerical solutions that plot infections versus time demonstrate a progression from zero effect to complete epidemic avoidance with a sharp drop-off, or knee, as the mask efficiency and mask adoption levels are increased to their calculated critical levels.

A fundamental reason why population-wide mask wearing should be effective is that, between two mask wearers, the concentration of particles at inhalation would be the square of the mask penetration fraction. Since squaring can be a much more powerful effect than a linear effect, some significant gains remain for less efficient masks and incomplete adoption by the population. The combination, or team, of masks can provide a strong dose-lowering squaring effect, which enable the use of lower-efficiency, lower-cost, lower pressure-drop (easier breathing) masks.

### 3.2. Limitations

Deterministic SIR models implicitly assume that the population has homogeneous mixing, with a fixed product of the number of contacts per day and of the probability of infection per contact. However, in some situations, for example at home, one would not normally expect people to wear masks. (Such limitations also apply to physical distancing.)

Since the basic reproduction number *R*_0_ is not a constant determined solely by viral characteristics, one might consider a range of *R*_0_ in a population in order to accommodate higher-density higher-contact regions. For example, regions with busy subways would be expected to have higher *R*_0._

The critical mask efficiency calculated herein may be insufficient in environments with exposure higher than the average expected in simple SIR modelling. One such environment would be hospitals. Another such environment would be long-term-care facilities, where the duration and closeness of the contacts between personal care workers and residents are significant. Additionally, mask wearing by residents would not be practical, so the advantages of combined mask wearing would not exist. Finally, such residents have a high death rate once infected. Given all of these factors, higher efficiency masks worn by personal care workers deserves consideration.

Other situations of higher-exposure than average might include those with tight-spacing and/or longer-duration, including public transportation and air travel.

Many situations may have average exposure, including shoppers in grocery stores and street-front retail shops.

### 4.3. Some Practical Mask Considerations

In the example of *R*_0_ = 2.5 epidemics, while surgical masks already surpass the critical mask efficiency, mask leakage can be improved by using an overlay of a nylon stocking (Cooper et al., 1983; Mueller and Fernandez, 2020). As reported in (Mueller and Fernandez, 2020) the use of a nylon stocking overlay raised the efficiency of several fabric masks above that of a benchmark surgical mask. Mimicking that approach, perhaps by increasing elasticity near the borders of masks, might significantly improve fit and performance of alternatives to surgical masks. Promising results using hybrid combinations of common fabrics is reported in (Konda et al., 2020).

Re-use of masks in pandemic situations is a natural consideration (Institute of Medicine, 2006), not just for low supply but also to lower costs. Some possible approaches include delayed re-use and disinfection. In (Pastorino et al., 2020) a million-fold inactivation of SARS-CoV-2 on N95 masks was achieved using 15 minutes of dry heat at 92C. In (Zulauf et al., 2020) a microwave-oven steam treatment for 3 minutes attained similar results. In (Lin et al., 2017) a gauze-type surgical mask retained good filtering efficiency of about 0.78, at its most-penetrating particle size, after decontamination using 3 minutes of dry heat at roughly 155 C.

## Data Availability

No data is required for independent verification.

## Appendix A Effective Reproduction Number

Here we determine the effective reproduction number *R*_0,*e*_ for the compartmental model of Figure 2 with its accompanying equations. We use the method of (van den Driessche and Watmough, 2002). Let

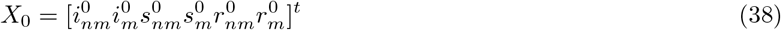

denote the state vector of our model at a disease free equilibrium (DFE). A DFE is a state in which the population remains without disease, so it will have 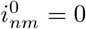 and 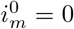 Additionally, we set 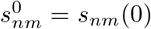 and 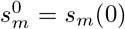 in order to determine conditions for which the initial conditions will progress to being disease free.

Guided by (van den Driessche and Watmough, 2002), the relevant matrices for our set of equations, based on our set of equations for the model, are;

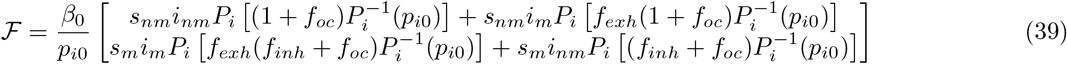

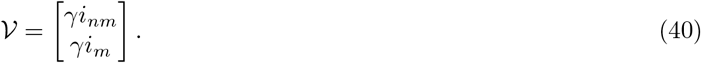

Then

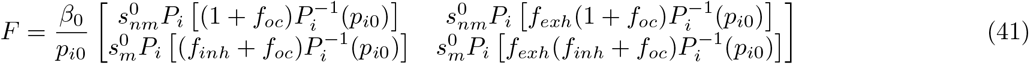

and

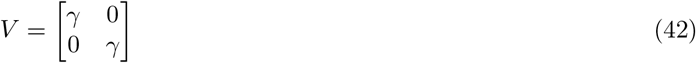

so that

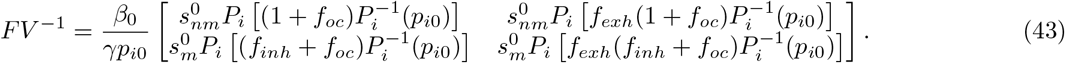

The largest eigenvalue of *FV* ^−1^ gives *R*_0,*e*_,

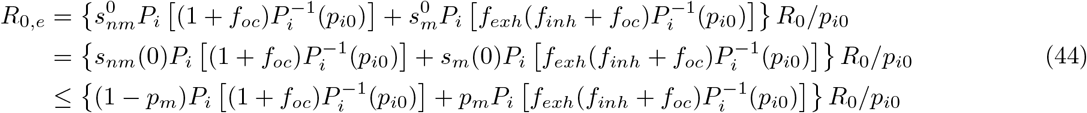

where *R*_0_ = *β*_0_/*γ*, and where, since *s*_*nm*_(0) = 1 − *p*_*m*_ − *i*_*nm*_(0) ≤ 1 − *p*_*m*_ and *s*_*m*_(0) = *p*_*m*_ − *i*_*m*_(0) ≤ *p*_*m*_, we have conservatively neglected the typically very small initial infectious fractions *i*_*nm*_(0), *i*_*m*_(0).

There are some possible simplifications to (44). If the masks have equal filter transmissions *f*_*t*_ then

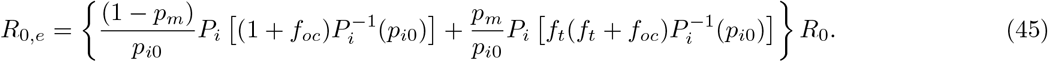

Additionally, if the contribution from the ocular route is not significant, then

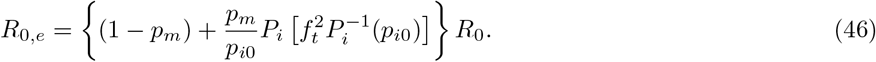

## Appendix B Effective *R*_0_ for Various Dose Response Functions

Here for each of the linear, exponential, and approximate Beta-Poisson dose-response functions, we use their inverses to give their response after dose scaling from mask filtering, and then use that result in the general expression (44) from Appendix A for *R*_0_*e*.

All of the dose-response functions below are setup for normalized doses, i.e. a dose *d* = 1 is the median-effective dose for a population, so that *P*_*i*_(1) = 0.5.

### B.1. Linear Dose Response

The linear dose-response is

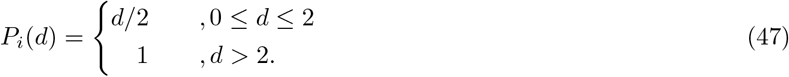

Its inverse is 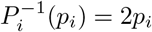 and the response after scaling a dose corresponding to an initial *P*_*i*0_ is

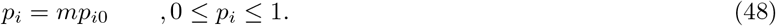

Using *p*_*i*_ in (44) we obtain

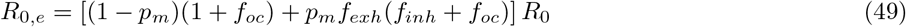

and for identical filter transmission gains *f*_*t*_ then

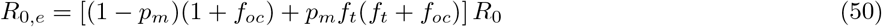

and with no ocular contribution,

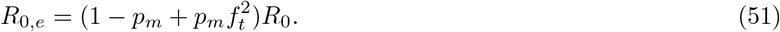

### B.2. Exponential Dose Response

The exponential dose-response is

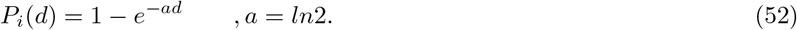

Its inverse is

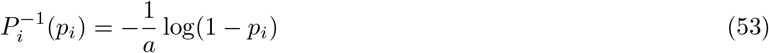

and the response after scaling a dose corresponding to an initial *P*_*i*0_ is

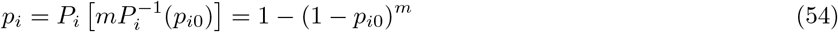

Using *p*_*i*_ in (44) we obtain

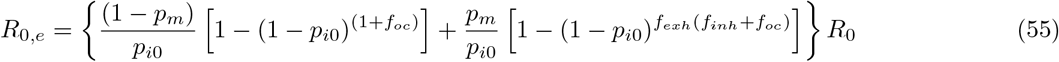

With identical filter transmission gains *f*_*t*_ then

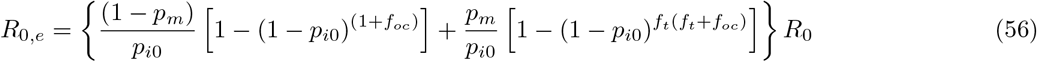

and with no ocular contribution,

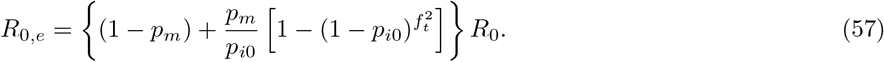

### B.3. Approximate Beta-Poisson Dose Response

The approximate beta-Poisson dose-response is

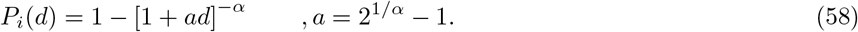

Its inverse is

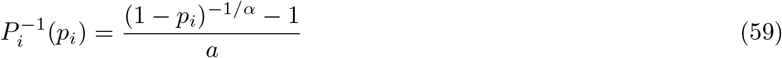

and the response after scaling a dose corresponding to an initial *P*_*i*0_ is

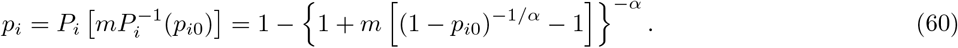

Using *p*_*i*_ in (44) we obtain

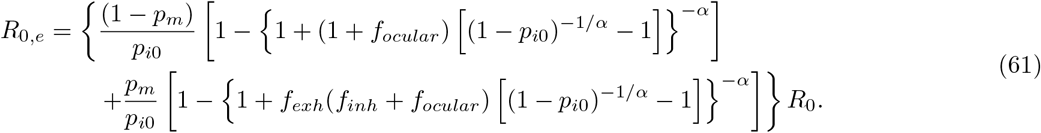

With identical filter transmission gains *f*_*t*_, then

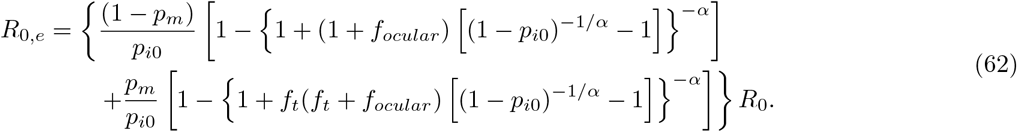

and with no ocular contribution,

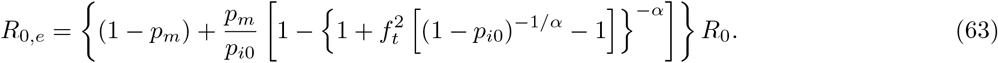

## Appendix C Critical Value of Mask Efficiency

Here for each of the linear, exponential, and approximate Beta-Poisson dose-response functions, we use the corresponding *R*_0,*e*_ determined in Appendix B, set it to unity and solve for the critical value of the mask efficiency. We assume that the masks have identical filter transmission gains *f*_*t*_.

### C.1. Linear Dose Response

With a linear dose response, setting *R*_0,*e*_ = 1 in (49) gives the condition,

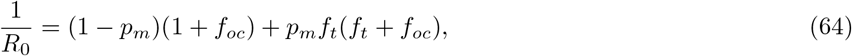

which can be solved to give the critical value of *f*_*t*_, or the critical mask efficiency *f*_*b,crit*_ = 1 − *f*_*t,crit*_ as,

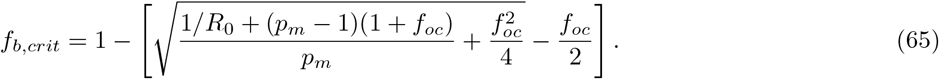

For no ocular contribution this simplifies to

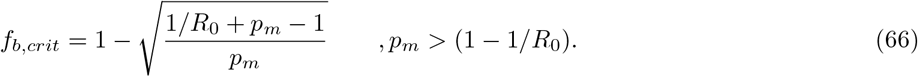

### C.2. Exponential Dose Response

With an exponential dose response, and neglecting the contribution from the ocular route, setting *R*_0,*e*_ = 1 in (57) gives the condition,

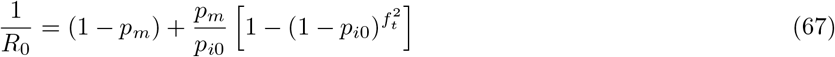

or

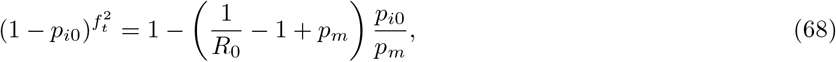

which can be solved to give the critical value of *f*_*t*_ or the critical value of mask efficiency *f*_*b,crit*_ = 1 − *f*_*t,crit*_ as,

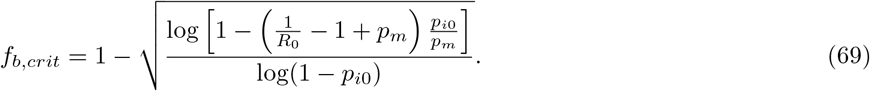

### C.3. Approximate Beta-Poisson Response

With an approximate beta-Poisson dose response, and neglecting the contribution from the ocular route, setting *R*_0,*e*_ = 1 in (63) gives the condition,

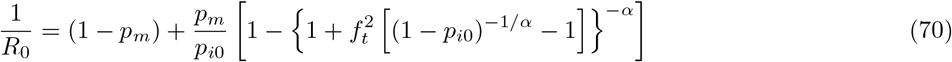

or

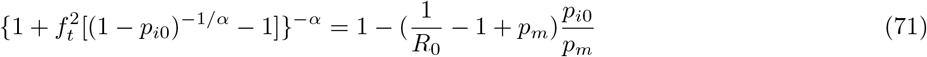

which can be solved to give the critical value of *f*_*t*_ or the critical value of mask efficiency *f*_*b,crit*_ = 1 − *f*_*t,crit*_ as,

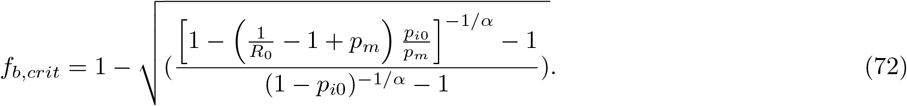

## Declaration of competing interests

None.

## Notes

### Competing Interest Statement

The authors have declared no competing interest.

### Funding Statement

No funding was received.

## References

1. Anfinrud, P., Stadnytskyi, V., Bax, C.E., Bax, A., 2020. Visualizing Speech-Generated Oral Fluid Droplets with Laser Light Scattering. New England Journal of Medicine, NEJMc2007800 doi:10.1056/NEJMc2007800.

2. Atkinson, M.P., Wein, L.M., 2008. Quantifying the Routes of Transmission for Pandemic Influenza. Bulletin of Mathematical Biology 70, 820–867. doi:10.1007/s11538-007-9281-2.

3. Bischoff, W.E., Reid, T., Russell, G.B., Peters, T.R., 2011. Transocular Entry of Seasonal Influenza-Attenuated Virus Aerosols and the Efficacy of N95 Respirators, Surgical Masks, and Eye Protection in Humans. Journal of Infectious Diseases 204, 193–199. doi:10.1093/infdis/jir238.

4. Bourouiba, L., 2020. Turbulent Gas Clouds and Respiratory Pathogen Emissions: Potential Implications for Reducing Transmission of COVID-19. JAMA doi:10.1001/jama.2020.4756.

5. Brienen, N.C.J., Timen, A., Wallinga, J., Van Steenbergen, J.E., Teunis, P.F.M., 2010. The Effect of Mask Use on the Spread of Influenza During a Pandemic: The Effect of Mask Use on the Spread of Influenza During a Pandemic. Risk Analysis 30, 1210–1218. doi:10.1111/j.1539-6924.2010.01428.x.

6. Brouwer, A.F., Weir, M.H., Eisenberg, M.C., Meza, R., Eisenberg, J.N.S., 2017. Dose-response relationships for environmentally mediated infectious disease transmission models. PLOS Computational Biology 13, e1005481. doi:10.1371/journal.pcbi.1005481.

7. Coburn, B.J., Wagner, B.G., Blower, S., 2009. Modeling influenza epidemics and pandemics: Insights into the future of swine flu (H1N1). BMC Medicine 7, 30. doi:10.1186/1741-7015-7-30.

8. Cooper, D.W., Hinds, W.C., Price, J.M., Weker, R., Yee, H.S., 1983. Common Materials for Emergency Respiratory Protection: Leakage Tests with a Manikin. American Industrial Hygiene Association Journal 44, 720–726. doi:10.1080/15298668391405634.

9. Cui, J., Zhang, Y., Feng, Z., Guo, S., Zhang, Y., 2019. Influence of asymptomatic infections for the effectiveness of facemasks during pandemic influenza. Mathematical Biosciences and Engineering 16, 3936–3946. doi:10.3934/mbe.2019194.

10. Davies, A., Thompson, K.A., Giri, K., Kafatos, G., Walker, J., Bennett, A., 2013. Testing the Efficacy of Homemade Masks: Would They Protect in an Influenza Pandemic? Disaster Medicine and Public Health Preparedness 7, 413–418. doi:10.1017/dmp.2013.43.

11. Eikenberry, S.E., Mancuso, M., Iboi, E., Phan, T., Eikenberry, K., Kuang, Y., Kostelich, E., Gumel, A.B., 2020. To mask or not to mask: Modeling the potential for face mask use by the general public to curtail the COVID-19 pandemic. Infectious Disease Modelling 5, 293–308. doi:10.1016/j.idm.2020.04.001.

12. Fears, A.C., Klimstra, W.B., Duprex, P., Hartman, A., Weaver, S.C., Plante, K.S., Mirchandani, D., Plante, J., Aguilar, P.V., Fernandez, D., Nalca, A., Totura, A., Dyer, D., Kearney, B., Lackemeyer, M., Bohannon, J.K., Johnson, R., Garry, R.F., Reed, D.S., Roy, C.J., 2020. Comparative Dynamic Aerosol Efficiencies of Three Emergent Coronaviruses and the Unusual Persistence of SARS-CoV-2 in Aerosol Suspensions. Preprint. Infectious Diseases (except HIV/AIDS). doi:10.1101/2020.04.13.20063784.

13. Ferguson, N., Cummings, D.A.T., Fraser, C., Cajka, J.C., Cooley, P.C., Burke, D.S., 2006. Strategies for mitigating an influenza pandemic. Nature 442, 448–452. doi:10.1038/nature04795.

14. Ferguson, N., Laydon, D., Nedjati Gilani, G., Imai, N., Ainslie, K., Baguelin, M., Bhatia, S., Boonyasiri, A., Cucunuba Perez, Z., Cuomo-Dannenburg, G., Dighe, A., Dorigatti, I., Fu, H., Gaythorpe, K., Green, W., Hamlet, A., Hinsley, W., Okell, L., Van Elsland, S., Thompson, H., Verity, R., Volz, E., Wang, H., Wang, Y., Walker, P., Winskill, P., Whittaker, C., Donnelly, C., Riley, S., Ghani, A., 2020. Report 9: Impact of Non-Pharmaceutical Interventions (NPIs) to Reduce COVID19 Mortality and Healthcare Demand. Technical Report. Imperial College London. doi:10.25561/77482.

15. Hethcote, H.W., 2000. The Mathematics of Infectious Diseases. SIAM Review 42, 599–653. doi:10.1137/S0036144500371907.

16. Institute of Medicine, 2006. Reusability of Facemasks During an Influenza Pandemic: Facing the Flu. National Academies Press, Washington, D.C. doi:10.17226/11637.

17. Jung, H., Kim, J.K., Lee, S., Lee, J., Kim, J., Tsai, P., Yoon, C., 2014. Comparison of Filtration Efficiency and Pressure Drop in Anti-Yellow Sand Masks, Quarantine Masks, Medical Masks, General Masks, and Handkerchiefs. Aerosol and Air Quality Research 14, 991–1002. doi:10.4209/aaqr.2013.06.0201.

18. Konda, A., Prakash, A., Moss, G.A., Schmoldt, M., Grant, G.D., Guha, S., 2020. Aerosol Filtration Efficiency of Common Fabrics Used in Respiratory Cloth Masks. ACS Nano, acsnano.0c03252 doi:10.1021/acsnano.0c03252.

19. Lau, J.T., Tsui, H., Lau, M., Yang, X., 2004. SARS Transmission, Risk Factors, and Prevention in Hong Kong. Emerging Infectious Diseases 10, 587–592. doi:10.3201/eid1004.030628.

20. Lee, S.A., Grinshpun, S., Reponen, T., 2008. Respiratory Performance Offered by N95 Respirators and Surgical Masks: Human Subject Evaluation with NaCl Aerosol Representing Bacterial and Viral Particle Size Range. The Annals of Occupational Hygiene doi:10.1093/annhyg/men005.

21. Leung, N.H.L., Chu, D.K.W., Shiu, E.Y.C., Chan, K.H., McDevitt, J.J., Hau, B.J.P., Yen, H.L., Li, Y., Ip, D.K.M.,Peiris, J.S.M., Seto, W.H., Leung, G.M., Milton, D.K., Cowling, B.J., 2020. Respiratory virus shedding in exhaled breath and efficacy of face masks. Nature Medicine doi:10.1038/s41591-020-0843-2.

22. Lin, T.H., Chen, C.C., Huang, S.H., Kuo, C.W., Lai, C.Y., Lin, W.Y., 2017. Filter quality of electret masks in filtering 14.6–594 nm aerosol particles: Effects of five decontamination methods. PLOS ONE 12, e0186217. doi:10.1371/journal.pone.0186217.

23. Milton, D.K., Fabian, M.P., Cowling, B.J., Grantham, M.L., McDevitt, J.J., 2013. Influenza Virus Aerosols in Human Exhaled Breath: Particle Size, Culturability, and Effect of Surgical Masks. PLoS Pathogens 9, e1003205. doi:10.1371/journal.ppat.1003205.

24. Mossong, J., Hens, N., Jit, M., Beutels, P., Auranen, K., Mikolajczyk, R., Massari, M., Salmaso, S., Tomba, G.S., Wallinga, J., Heijne, J., Sadkowska-Todys, M., Rosinska, M., Edmunds, W.J., 2008. Social Contacts and Mixing Patterns Relevant to the Spread of Infectious Diseases. PLoS Medicine 5, e74. doi:10.1371/journal.pmed.0050074.

25. Mueller, A.V., Fernandez, L.A., 2020. Assessment of Fabric Masks as Alternatives to Standard Surgical Masks in Terms of Particle Filtration Efficiency. Preprint. Occupational and Environmental Health. doi:10.1101/2020.04.17.20069567.

26. Murphy, B.R., Clements, M.L., Madore, H.P., Steinberg, J., O’Donnell, S., Betts, R., Demico, D., Reichman, R.C., Dolin, R., Maassab, H.F., 1984. Dose Response of Cold-Adapted, Reassortant Influenza A/California/10/78 Virus (H1N1) in Adult Volunteers. Journal of Infectious Diseases 149, 816–816. doi:10.1093/infdis/149.5.816.

27. Myers, M.R., Hariharan, P., Guha, S., Yan, J., 2016. A mathematical model for assessing the effectiveness of protective devices in reducing risk of infection by inhalable droplets. Mathematical Medicine and Biology, dqw018 doi:10.1093/imammb/dqw018.

28. Ngonghala, C.N., Iboi, E., Eikenberry, S., Scotch, M., MacIntyre, C.R., Bonds, M.H., Gumel, A.B., 2020. Mathematical assessment of the impact of non-pharmaceutical interventions on curtailing the 2019 novel Coronavirus. Mathematical Biosciences 325, 108364. doi:10.1016/j.mbs.2020.108364.

29. Oberg, T., Brosseau, L.M., 2008. Surgical mask filter and fit performance. American Journal of Infection Control 36, 276–282. doi:10.1016/j.ajic.2007.07.008.

30. Pastorino, B., Touret, F., Gilles, M., de Lamballerie, X., Charrel, R.N., 2020. Evaluation of Heating and Chemical Protocols for Inactivating SARS-CoV-2. Preprint. Microbiology. doi:10.1101/2020.04.11.036855.

31. Rengasamy, S., Eimer, B., Shaffer, R., 2010. Simple Respiratory Protection—Evaluation of the Filtration Performance of Cloth Masks and Common Fabric Materials Against 20–1000 nm Size Particles. The Annals of Occupational Hygiene doi:10.1093/annhyg/meq044.

32. Smith, J.D., MacDougall, C.C., Johnstone, J., Copes, R.A., Schwartz, B., Garber, G.E., 2016. Effectiveness of N95 respirators versus surgical masks in protecting health care workers from acute respiratory infection: A systematic review and meta-analysis. Canadian Medical Association Journal 188, 567–574. doi:10.1503/cmaj.150835.

33. Stadnytskyi, V., Bax, C.E., Bax, A., Anfinrud, P., 2020. The airborne lifetime of small speech droplets and their potential importance in SARS-CoV-2 transmission. Proceedings of the National Academy of Sciences, 202006874 doi:10.1073/pnas.2006874117.

34. Stilianakis, N.I., Drossinos, Y., 2010. Dynamics of infectious disease transmission by inhalable respiratory droplets. Journal of The Royal Society Interface 7, 1355–1366. doi:10.1098/rsif.2010.0026.

35. Taylor, C., Kampf, S., Grundig, T., 2020. Taiwan is beating COVID-19 without closing schools or workplaces. Can Canada do the same? — CBC News. https://www.cbc.ca/news/business/taiwan-covid-19-lessons-1.5505031.

36. Tracht, S.M., Del Valle, S.Y., Hyman, J.M., 2010. Mathematical Modeling of the Effectiveness of Facemasks in Reducing the Spread of Novel Influenza A (H1N1). PLoS ONE 5, e9018. doi:10.1371/journal.pone.0009018.

37. van den Driessche, P., Watmough, J., 2002. Reproduction numbers and sub-threshold endemic equilibria for compartmental models of disease transmission. Mathematical Biosciences 180, 29–48. doi:10.1016/S0025-5564(02)00108-6.

38. van der Sande, M., Teunis, P., Sabel, R., 2008. Professional and Home-Made Face Masks Reduce Exposure to Respiratory Infections among the General Population. PLoS ONE 3, e2618. doi:10.1371/journal.pone.0002618.

39. van Doremalen, N., Bushmaker, T., Morris, D.H., Holbrook, M.G., Gamble, A., Williamson, B.N., Tamin, A., Harcourt, J.L., Thornburg, N.J., Gerber, S.I., Lloyd-Smith, J.O., de Wit, E., Munster, V.J., 2020. Aerosol and Surface Stability of SARS-CoV-2 as Compared with SARS-CoV-1. New England Journal of Medicine 382, 1564–1567. doi:10.1056/NEJMc2004973.

40. Watanabe, T., Bartrand, T.A., Weir, M.H., Omura, T., Haas, C.N., 2010. Development of a Dose-Response Model for SARS Coronavirus: Dose-Response Model for SARS-CoV. Risk Analysis 30, 1129–1138. doi:10.1111/j.1539-6924.2010.01427.x.

41. Wei, J., Li, Y., 2016. Airborne spread of infectious agents in the indoor environment. American Journal of Infection Control 44, S102–S108. doi:10.1016/j.ajic.2016.06.003.

42. Wei, W.E., Li, Z., Chiew, C.J., Yong, S.E., Toh, M.P., Lee, V.J., 2020. Presymptomatic Transmission of SARS-CoV-2 — Singapore, January 23–March 16, 2020. MMWR. Morbidity and Mortality Weekly Report 69, 411–415. doi:10.15585/mmwr.mm6914e1.

43. Wein, L.M., Atkinson, M.P., 2009. Assessing Infection Control Measures for Pandemic Influenza. Risk Analysis 29, 949–962. doi:10.1111/j.1539-6924.2009.01232.x.

44. Xie, X., Li, Y., Chwang, A.T.Y., Ho, P.L., Seto, W.H., 2007. How far droplets can move in indoor environments, revisiting the Wells evaporation falling curve. Indoor Air 17, 211–225. doi:10.1111/j.1600-0668.2007.00469.x.

45. Yan, J., Guha, S., Hariharan, P., Myers, M., 2019. Modeling the Effectiveness of Respiratory Protective Devices in Reducing Influenza Outbreak: Modeling the Effectiveness of Respiratory Protective Devices. Risk Analysis 39, 647–661. doi:10.1111/risa.13181.

46. Zulauf, K.E., Green, A.B., Ba, A.N.N., Jagdish, T., Reif, D., Seeley, R., Dale, A., Kirby, J.E., 2020. Microwave-Generated Steam Decontamination of N95 Respirators Utilizing Universally Accessible Materials. Preprint. Occupational and Environmental Health. doi:10.1101/2020.04.22.20076117.

